# New manual qPCR assay validated on tongue swabs collected and processed in Uganda shows sensitivity that rivals sputum-based molecular TB diagnostics

**DOI:** 10.1101/2023.08.10.23293680

**Authors:** Amy Steadman, Alfred Andama, Alexey Ball, Job Mukwatamundu, Khushboo Khimani, Tessa Mochizuki, Lucy Asege, Alice Bukirwa, John Baptist Kato, David Katumba, Esther Kisakye, Wilson Mangeni, Sandra Mwebe, Martha Nakaye, Irene Nasuna, Justine Nyawere, Deryk Visente, Catherine Cook, Talemwa Nalugwa, Christine M. Bachman, Fred Semitalia, Bernhard H. Weigl, John Connelly, William Worodria, Adithya Cattamanchi

## Abstract

**Background:** Reliance on sputum-based testing is a key barrier to increasing access to molecular diagnostics for tuberculosis (TB). Many people with TB are unable to produce and sputum processing increases the complexity and cost of molecular assays. Tongue swabs are emerging as an alternative to sputum, but performance limits are uncertain.

**Methods:** From June 2022 to July 2023, we enrolled 397 consecutive adults with cough >2 weeks at two health centers in Kampala, Uganda. We collected routine demographic and clinical information, sputum for routine TB testing (one Xpert MTB/RIF Ultra® and two liquid cultures), and up to four tongue swabs for same-day qPCR. We evaluated tongue swab qPCR diagnostic accuracy in reference to sputum TB test results, quantified TB targets per swab, assessed the impact of serial swabbing, and compared two swab types (Copan FLOQSWAB® and Steripack® spun polyester swabs).

**Results:** Among 397 participants, 43.1% were female, median age was 33 years, 23.5% were living with HIV (PLHIV) and 32.3% had confirmed TB. Sputum Xpert Ultra and tongue swab qPCR results were concordant for 98.2% [96.2-99.1] of participants. Tongue swab qPCR sensitivity was 91.0% [84.6-94.9] and specificity 98.9% [96.2-99.8] vs. microbiological reference standard (MRS). A single tongue swab recovered a seven-log range of TB copies, with a decreasing recovery trend among four serial swabs. We found no difference between swab types.

**Conclusions:** Tongue swabs show promise as an alternative to sputum for TB diagnosis, with sensitivity approaching sputum-based molecular tests. Our results provide valuable insights for developing successful tongue swab-based TB diagnostics.

## INTRODUCTION

Tuberculosis was responsible for 1.6 million deaths in 2021, with disruptions to diagnosis and treatment caused by the COVID-19 pandemic. TB is curable and preventable, but diagnosis remains the largest gap in the care cascade with the number of new notifications already falling 18% from 2019 to 2020, and missed diagnosis for an additional 4.2 million during the pandemic, highlighting the urgent need for improved access to diagnostic services^1-2^.

Currently, many of the TB diagnostics available in the world’s highest-burden countries rely on high-quality sputum, which is a challenging specimen type for several reasons: Children, people living with HIV (PLHIV), and others lacking productive cough are often unable to provide quality samples^3,4^. Sputum collection may release infectious bioaerosols, creating risks to health care workers and nearby patients^5–10^, resulting in reluctance to order sputum-based tests^11^. Further, laboratory-based diagnostics, such as sputum smear microscopy, have suboptimal diagnostic accuracy, while sputum culture’s long turnaround time (TAT) often results in missed or delayed treatment^4,12,13^.

The World Health Organization (WHO) has recommended molecular diagnostics like GeneXpert® MTB/RIF Ultra (Cepheid) and Truenat® MTB Plus (Molbio) due to improved accuracy and shorter TAT, but barriers to access remain. In Uganda, the estimated cost per patient tested with Xpert Ultra is over $21.00 USD, 80% of which is due to the equipment and cartridge costs^8,14^.

Oral swabbing has recently been shown to be a compelling alternative to sputum collection that may increase access to molecular diagnostics for TB^15^. The COVID-19 pandemic highlighted the acceptability and efficacy of swab-based approaches, because they are inexpensive, amenable to self-collection, and noninvasive^16^. Further, the reduced complexity of oral matrix compared to sputum makes it possible to employ extraction-free sample prep methods, reducing TAT and the need for additional consumables and equipment.

Publications on tongue swab clinical studies, methods and outcomes metrics, including sensitivity and specificity, have varied^15,17–23^, but they hint at keys to increasing performance of swab-based testing. For instance, it has been shown that MTB-containing biofilms form on tongue papillae, with higher MTB recovery reported from the tongue compared to other oral sites (cheek, gum)^18,24^.

We hypothesized that we may enhance the sensitivity of tongue-swab-based molecular tests by optimizing several assay components, while revealing the quantity of MTB that can be recovered from the tongue. There are opportunities for technical gains in sample prep, because MTB is resistant to conventional bacterial lysis techniques due to the complex structure of its cell envelope, which is comprised of lipophilic molecules including long-chain mycolic acids and polysaccharides^25^. Low lysis efficiency may yield artificially low sensitivity, and its optimization may increase detection of MTB^26^. While other studies employed DNA concentration and purification of tongue swab specimens^15^, we discovered this may be a source of recovery losses. To circumvent these steps, we determined that downstream amplification and detection techniques must be inhibitor-tolerant, and input volumes must be high enough to limit stochastic sampling error^27^.

Past publications have highlighted the consistent presence of total bacterial biomass on 10 serially collected tongue swabs^20^, and we similarly evaluated MTB-specific depletion after collection of four serial swabs. Our findings elucidate the total amount of MTB on the tongue and underscore the potential utility of novel sampling tools. We compared the recovery of the Copan FLOQSWAB® and Steripack® spun polyester swabs to understand the limitations of currently available sampling options.

The present study validated a novel quantitative triplex qPCR assay for TB on tongue swabs collected and processed within 24 hours. Our results quantify TB targets on the tongue for the first time and demonstrate that increased efficiency of collection, lysis, and amplification and detection achieve high concordance with sputum Xpert Ultra testing, even in the absence of DNA extraction. These methods provide benchmarks for tongue swab assay development and may serve as a roadmap for TB diagnostics manufacturers.

## METHODS

### Study participants

We enrolled consenting adults (>12-years-old) presenting with at least two weeks cough to two health centers in Kampala, Uganda between June 28, 2022 and July 24, 2023. We excluded people who had been treated for TB infection or disease in the last 12 months, had taken antibiotics with antimycobacterial activity in the last two weeks, or were unable or unwilling to return for follow-up or provide informed consent.

The study was approved by the Makerere University School of Medicine Research and Ethics Committee (2020-182) and the Ugandan National Council on Science and Technology (HS1482ES). Clinical and laboratory staff were blinded to TB status during collection and processing. Participants were assigned identification numbers with the prefix “R2D204…” which were not known to anyone outside the research team.

### Procedures

We collected detailed demographic, TB symptom and medical history using a standardized case report form. All participants had finger prick or venous blood collected for HIV testing. Up to three spot sputum samples were collected for reference standard testing, which included Xpert Ultra (with repeat testing if the initial result was Trace-positive, invalid, or indeterminate) and two cultures in liquid Mycobacterium Growth Indicator Tube (MGIT) media. Prior to sputum collection, up to four tongue swabs were collected from each participant. Throughout three sub-studies, the first swab was always a Copan FLOQSWAB® (520CS01) to ensure a common thread for diagnostic accuracy and copy number calculations. Firm pressure was applied to the swab handle while swabbing the entire length and breadth of the anterior three quarters of the tongue dorsum for 30 seconds to ensure sampling uniformity. Swab timing was shortened to 15 seconds with a focus on the posterior portion of the anterior three quarters of the tongue dorsum for sub-study 3. After completion, the swab head was inserted into the top of a gasketed screw cap tube containing 500μl 1X Tris-EDTA (TE) pH 7.4 or pH 8.0 (preferred) and broken at the 30mm breakpoint. Tubes were labeled to indicate swab order and stored in a cooler box containing ice packs until transportation to the laboratory for same-day analysis.

For the serial swabbing sub-study, three additional FLOQSWAB^®^ were sequentially collected (four identical swabs per participant). For the swab comparison sub-study, two additional swabs were collected. The first swab was always a whole-tongue FLOQSWAB^®^ as described above. Afterward, two half-tongue swabs were collected for 30 seconds each using the centerline of the tongue as a guide. One side of the tongue was swabbed with a FLOQSWAB^®^ and the other side with a Steripack^®^ swab (60564RevC). Collection order was alternated daily.

Process control swabs were taken at each site once per week and processed along with clinical samples to ensure there was no contamination of the clinical workspaces or introduced during lab processing.

### Index test

#### Sample preparation

Tongue swab samples were vortexed for 15 seconds then immediately heated at 95°C for 30 minutes, vortexed again for 15 seconds, and centrifuged for 3 seconds. Maximum sample volume (approximately 375μl) was sterilely transferred to flat-bottom tubes (VWR 76417-214) containing 150mg of 0.1mm glass beads (RPI Corp 9830). Tubes were placed in a bead beater (BioSpec^®^ 607EUR), balanced crosswise, and subjected to three, one-minute beating cycles with one-minute rests in between. Tubes were centrifuged for 3 seconds and 320μl were removed into a fresh tube for qPCR.

#### Bioinformatics Analysis

Oligonucleotide sequences were generated in Geneious Prime^®^ version 2020.0.3 and screened for unfavorable folding and oligomerization using AutoDimer Version 1.022 with the following parameters: Minimum SCORE Requirement: 3; Na+ 0.085M; temp for dG calc 37°C; total strand conc 1.0μM. Sequences were screened for specificity to the *Mycobacterium tuberculosis* complex (MTBC) with NCBI Blast blastn.

#### qPCR

Five, 50μl aliquots crude lysate per sample were added to a PCR plate containing 10X KAPA3G^®^ (Roche 09160914103) and oligos targeting an MTB-complex-specific 85 bp region of the IS6110 insertion sequence, a 90 bp region of the IS1081 insertion sequence, and a 65 bp region of the RNaseP human sample adequacy control **(Supplemental 1)**. No template controls (NTCs) were run on each plate. Samples were processed on a QuantStudio5 0.2ml block thermal cycler. Quantities of MTB insertion elements were interpolated from aggregated MTB H37Rv DNA (ATCC^®^ 25618DQ) standard curves. Samples were considered positive if any of the five wells were positive.

Full standard operating procedures may be found in **Supplemental 2**. Additional assay optimization methods, including TB H37Ra cell-line culture methods and contrived sample generation, may be found in **Supplemental 3**.

### Reference standard definitions

We used a composite MRS to define TB status. Participants were considered to have active TB if they had a positive sputum Xpert Ultra result (Very Low or higher) and were considered negative for active TB if they had a negative MGIT culture and negative Xpert result. Participants who did not meet either criterion had an indeterminate TB status.

We also considered a reference standard that included only sputum Xpert Ultra results (SXRS). Participants were considered to have active TB if they had a positive Xpert result (Very Low or higher) or two Trace-positive results. Participants TB-negative if they had a negative Xpert result, without any Trace-positive results. Participants who did not meet either criterion had an indeterminant TB status.

### Data Analysis

To assess the impact of serial swabbing on MTB recovery, we fit a linear two-level mixed effects model for repeated measures to account for recovery differences among participants (random effect), with swab number as a fixed effect. We included MRS-positive participants and excluded participants with MTB detected from fewer than four swabs. We calculated the mean MTB copy number from five technical replicates. We log-transformed mean MTB copy number to account for the highly skewed distribution of the data.

To evaluate the difference in MTB target recovery from Copan and Steripack swabs, we performed a ratio paired T test. To understand whether differences between swabs were a function of the abundance of MTB cells on the swabs, we also performed a Bland-Altman analysis which calculated the percent difference of the tests divided by mean MTB IS6110 target recovery. We included participants who had a positive qPCR test from both the Copan and Steripack half-tongue swabs. We calculated mean MTB copy number per swab by transforming the mean copy number calculated per 50μl well using the function Y = 10*Y.

We analyzed differences in tongue swab IS6110 recovery by Xpert Ultra semi-quantitative categories using a Brown-Forsythe and Welch ANOVA test to correct for unequal standard deviations among groups and used a Dunnett T3 test to correct for multiple comparisons given the small (<50) sample size per group. Analyses were performed using GraphPad Prism version 9.3.0. and Stata Version 17 (StataCorp, College Station, TX, USA).

## RESULTS

### Participant characteristics

Between June 28, 2022 and July 24, 2023, 397 participants meeting eligibility criteria were enrolled for all three parts of this study. Tongue swabs were not collected from 6 (1.5%) participants who presented to the clinic too late for same-day sample processing and 13 (3.3%) participants with invalid tongue swab qPCR results. Of the remaining participants, 43.1% were female, median age was XX, XX were PLHIV and XX had MRS-confirmed TB **(Table 1)**.

**Table 1.** Demographic and clinical characteristics of study participants.

|  |  | Overall | Serial Swabbing<br>(Sub-study 1) | Copan vs. Steripack<br>(Sub-study 2) | Shortened Swab Time<br>(Sub-study 3) |
| --- | --- | --- | --- | --- | --- |
| **Total with valid qPCR result** |  | 378 | 97 | 133 | 148 |
| **Sex at birth** | Male | 215 (56.9%) | 53 (54.6%) | 79 (59.4%) | 83 (56.1%) |
|  | Female | 163 (43.1%) | 44 (45.4%) | 54 (40.6%) | 65 (43.9%) |
| **Median age (IQR)** |  | 33 (26, 43) | 32 (26, 42) | 36 (28, 43) | 32 (25, 42) |
| **PLHIV** |  | 89 (23.5%) | 22 (22.7%) | 35 (26.3%) | 32 (21.6%) |
| **Prior TB** |  | 55 (14.6%) | 19 (19.6%) | 16 (12.0%) | 20 (13.5%) |
| **Xpert Ultra Sputum result** | TB Negative | 262 (69.3%) | 64 (66.0%) | 90 (67.7%) | 108 (73.0%) |
|  | TB Positive | 113 (30.0%) | 32 (33.0%) | 41 (30.8%) | 40 (27.0%) |
|  | Trace | 3 (0.7%) | 1 (1.0%) | 2 (1.5%) | 0 (0%) |
| **Xpert semi-quantitative grade** | Trace | 3 (0.7%) | 1 (1.0%) | 2 (1.5%) | 0 (0%) |
|  | Very low | 5 (1.3%) | 0 (0%) | 3 (2.3%) | 2 (1.4%) |
|  | Low | 36 (9.5%) | 6 (6.2%) | 9 (6.8%) | 11 (7.4%) |
|  | Medium | 33 (8.7%) | 12 (12.4%) | 11 (8.3%) | 10 (6.8%) |
|  | High | 49 (13.0%) | 14 (14.4%) | 18 (13.5%) | 17 (11.5%) |
| **Microbiologic reference standard** | TB Negative | 190 (50.3%) | 62 (63.9%) | 74 (55.6%) | 54 (36.5%) |
|  | TB Positive | 122 (32.3%) | 33 (34.0%) | 46 (34.6%) | 42 (28.4%) |
|  | Indeterminate | 24 (6.3%) | 2 (2.1%) | 13 (9.7%) | 6 (4.1%) |
|  | *Pending* | 46 (12.2%) | 0 (0%) | 0 (0%) | 46 (31.1%) |

### Diagnostic accuracy of tongue swab qPCR

Among the 379 participants with valid qPCR results, concordance between sputum Xpert Ultra and tongue swab qPCR was 98.7% [95% CI: 96.9, 99.4] when Xpert Ultra Trace results were excluded and 98.2% [96.2, 99.1] when Xpert Ultra trace results were included. Sensitivity was 91.0% [95% CI: 84.6-94.9] for tongue swab qPCR and 94.3% [95% CI: 88.6-97.2] for sputum Xpert Ultra compared to the MRS.

Among the 190 MGIT culture-negative participants, concordance between sputum Xpert Ultra and tongue swab qPCR was 99.5% [95% CI 97.1, 100.0]. There was 1 participant with negative sputum Xpert Ultra but positive tongue swab qPCR results and positive MGIT culture results. Specificity was 98.9% [95% CI: 96.2, 99.8] for tongue swab qPCR and 99.5% [95% CI 97.1, 100.0] for sputum Xpert Ultra.

When comparing against sputum Xpert Ultra results alone, sensitivity of tongue swab PCR was 98.2% [95% CI: 93.8, 99.7] and specificity 98.9% [95% CI: 96.7, 99.7]. When Xpert Ultra Trace positive were included, sensitivity decreased to 96.6% [95% CI: 91.5, 98.7] **(Table 2)**.

**Table 2.** Diagnostic accuracy of tongue swabs compared to Xpert Ultra and MRS.

|  | Samples with valid paired Xpert and qPCR results |  |  | Excluded qPCR |  |
| --- | --- | --- | --- | --- | --- |
|  | Xpert (sputum) | qPCR (tongue swab) | Percent Agreement [95% CI] | Invalid | Not Tested |
| **Total enrolled = 397** | **378** | **378** |  | **13 (3.3%)** | **6 (1.5%)** |
| Overall agreement (without Trace) | 375 | 370 | 98.7 [96.9, 99.4] |  |  |
| Overall agreement (with Trace) | 378 | 371 | 98.2 [96.2, 99.1] |  |  |
| Negative | 262 | 259 | 98.9 [96.7, 99.7] | 10 (3.8%) | 5 (1.2%) |
| Positive (excluding Trace) | 113 | 111 | 98.2 [93.8, 99.7] | 3 (2.7%) | 0.0% |
| Positive (including Trace) | 116 | 112 | 96.6 [91.5, 98.7] | 3 (2.7%) | 1 (0.09)% |
| **Xpert Ultra Semi-Quantitative** |  |  |  |  |  |
| Very low | 5 | 3 | 60.0 [23.1, 92.9] | 1 |  |
| Low | 26 | 26 | 100.0 [86.7, 100.0] |  |  |
| Medium | 33 | 33 | 100.0 [89.8, 100.0] | 1 |  |
| High | 49 | 49 | 100.0 [92.6, 100.0] | 1 |  |
| Trace | 3 | 1 | 33.3 [1.3, 69.9] |  | 1 |
|  | Samples with valid paired MRS and qPCR results [95% CI] |  |  | Excluded qPCR |  |
|  | MRS (sputum) | qPCR (tongue swab) | Agreement | Invalid | Not Tested |
| **Total with valid results and culture data** | **312** | **312** |  | **13 (3.9%)** | **6 (1.8%)** |
| MRS Negative (culture negative) | 190 | 188 | 98.9 [96.2-99.8] | 8 | 2 |
| MRS Positive (culture plus Xpert) | 122 | 111 | 91.0 [84.6-94.9] | 4 | 1 |
| **Total** | **312** | **299** | **95.8 [93.0-97.5]** |  |  |
| Indeterminate | 24 |  |  | 1 | 3 |
| Pending Culture Results | 46 |  |  |  |  |
|  | Number of samples with valid matched MRS, Xpert, qPCR Results |  |  | Excluded qPCR |  |
|  | MRS (sputum) | Xpert Ultra (sputum) | Agreement | Invalid | Not Tested |
| **Total with valid results and culture data** | **312** | **312** |  | **13 (3.9%)** | **6 (1.8%)** |
| MRS Negative (culture negative) | 190 | 189 | 99.5 [97.1-100.0] | 8 | 2 |
| MRS Positive (culture plus Xpert) | 122 | 115 | 94.3 [88.6-97.2] | 4 | 1 |
| **Total** | **312** | **303** | **97.1 [94.6-98.5]** |  |  |
| Indeterminate | 24 |  |  | 1 | 3 |
| Pending Culture Results | 46 |  |  |  |  |
|  | **Culture Positive** | **Culture Negative** | **No Culture Data** |  |  |
| **Xpert Ultra Trace** | **1** | **1** | **2** |  | **1** |

### Quantification of MTB copies recovered from a single tongue swab

Detectable quantities of MTB were present on 111 swabs 122 were positive by the MRS. The number of MTB IS6110 targets per swab spanned a seven-log range and correlated with GeneXpert semi-quantitative categories, an indicator of bacillary load **(Figure 1)**. Mean IS6110 copies observed from tongue swabs were 369844, 106841, 8592, and 5200 compared to High, Medium, Low, and Very Low sputum Gene Xpert Ultra semi-quantitative categories, respectively. There were no significant differences between means of High and Medium (p = 0.0908) or Low and Very Low categories (p = 0.9864), though the latter finding may be skewed by the low number of participants with a Very Low sputum result.

**Figure 1.**
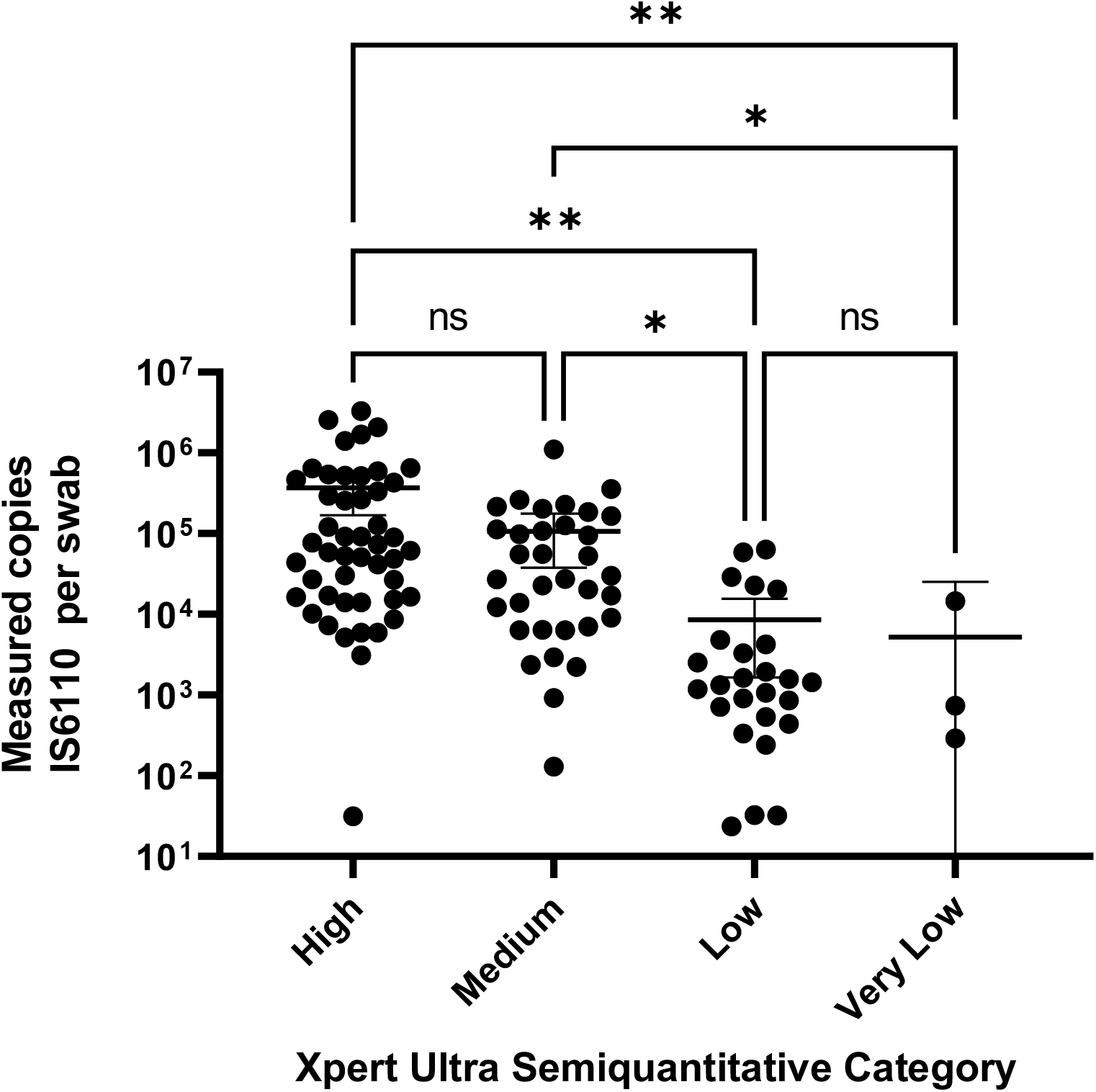
Measured mean log copies of MTB IS6110 per swab per participant, by Xpert semi-quantitative grade.

**Figure 2.**
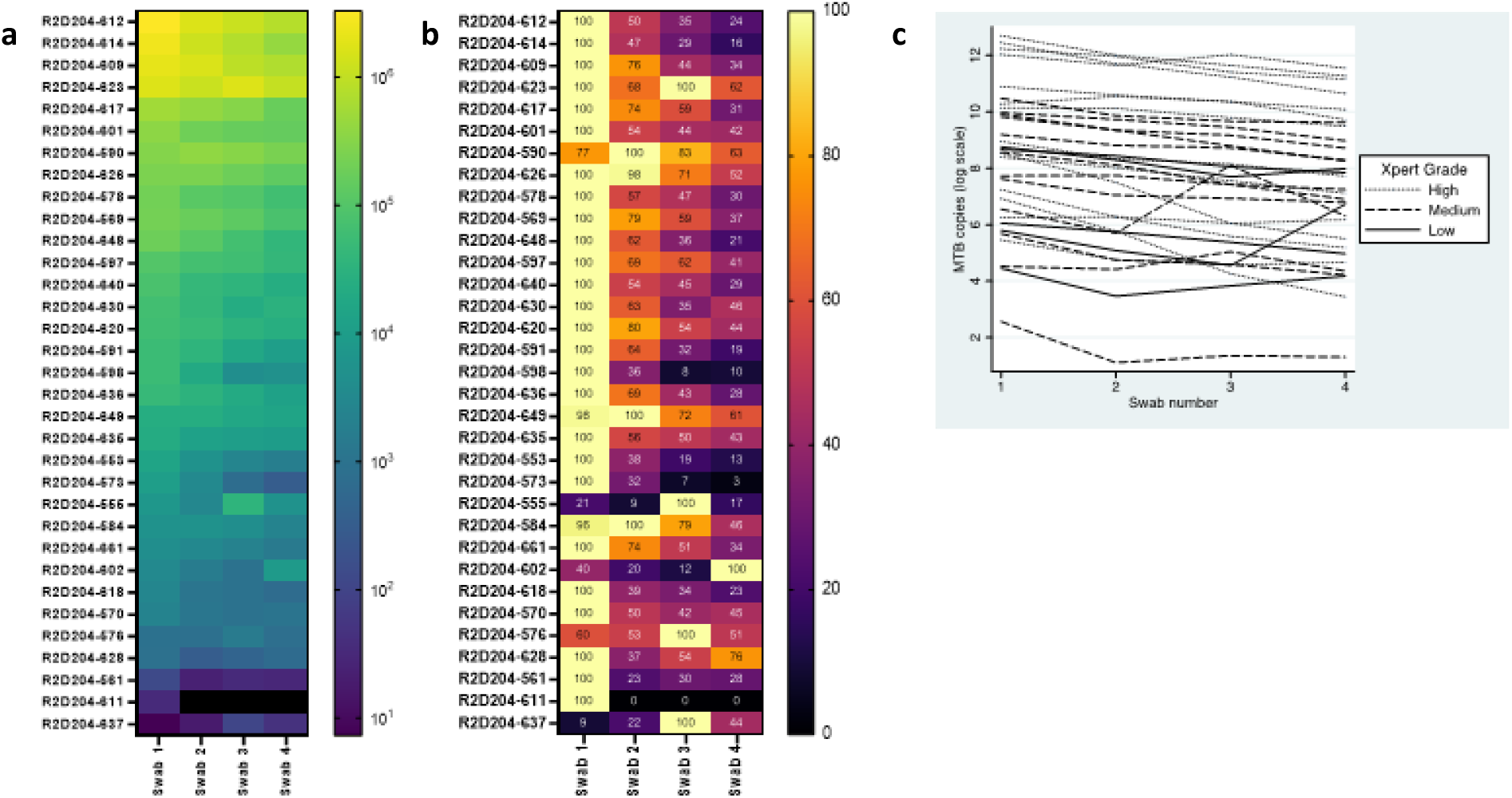
Quantity of MTB IS6110 copies present on four sequentially collected swabs. **a**. Heat map demonstrating decreasing recovery with each sequentially collected swab. **b**. Normalized MTB targets recovered from all swabs calculated as a percentage of the highest recovery condition (“100%”). **c**. Recovery of four swabs by Xpert semi-quantitative status.

### Serial swabbing

MTB was recovered from all four Copan swabs for 32 (100%) participants who were MRS-positive. We excluded one MRS-positive participant for whom MTB was only detected on the initial swab. We included one participant with a sputum Trace-positive Xpert Ultra result followed by a negative sputum Xpert Ultra result and a negative TB culture, because this participant was qPCR-positive on all four swabs.

Considering variation in MTB recovery across participants, we observed a decrease in MTB recovery with each sequential swab, when compared the initial swab (Swab 2 regression coefficient: -0.47 [95% CI: - 0.73, -0.22]; Swab 3 coefficient: -0.66 [95% CI: -0.91, -0.40]; Swab 4 coefficient: -0.96 [95% CI: -1.21, - 0.71]. Some variation was observed by Xpert semi-quantitative grade.

To confirm that MTB target identification was not due to differences in swabbing technique between swabs, we performed a delta delta CT calculation, enabling normalization to the RNaseP human gene. The delta delta CT also showed a decreasing trend of MTB yield by swab (**Supplement 2**).

### Copan and Steripack swab comparison

We recovered MTB from the first whole-tongue Copan swab taken from 39 (95.1%) MRS-positive participants. Of these, 37 participants had positive qPCR results for half-tongue Copan and Steripack swabs and were included in the analysis. Two participants were excluded because only one swab had a positive result. We observed two MRS-negative participants with weakly positive tongue swab results, and three participants were excluded due to negative process control contamination.

We used a ratio paired T-test to evaluate recovery from the two swab types due to large differences in bacillary load among participants. Copan log mean MTB recovery was 3.950 [95% CI: 3.555, 4.344] and Steripack log mean MTB recovery was 3.954 [95% CI: 3.600, 4.308]. The test indicated there is no statistically significant difference in MTB recovery between types (p = 0.9516), and the geometric mean of the ratios was 0.9902 and the pairing was significantly effective r=0.9354 (p = <0.0001). Normality of residuals was confirmed by Anderson-Darling (A2*), D’Agostino-Pearson omnibus (K2), Shapiro-Wilk (W), and Kolmogorov-Smirnov (distance) tests. A Bland-Altman analysis of the same data demonstrated no difference between Copan and Steripack recovery, regardless of the mean quantity of MTB targets per swab (**Figure 3)**.

**Figure 3.**
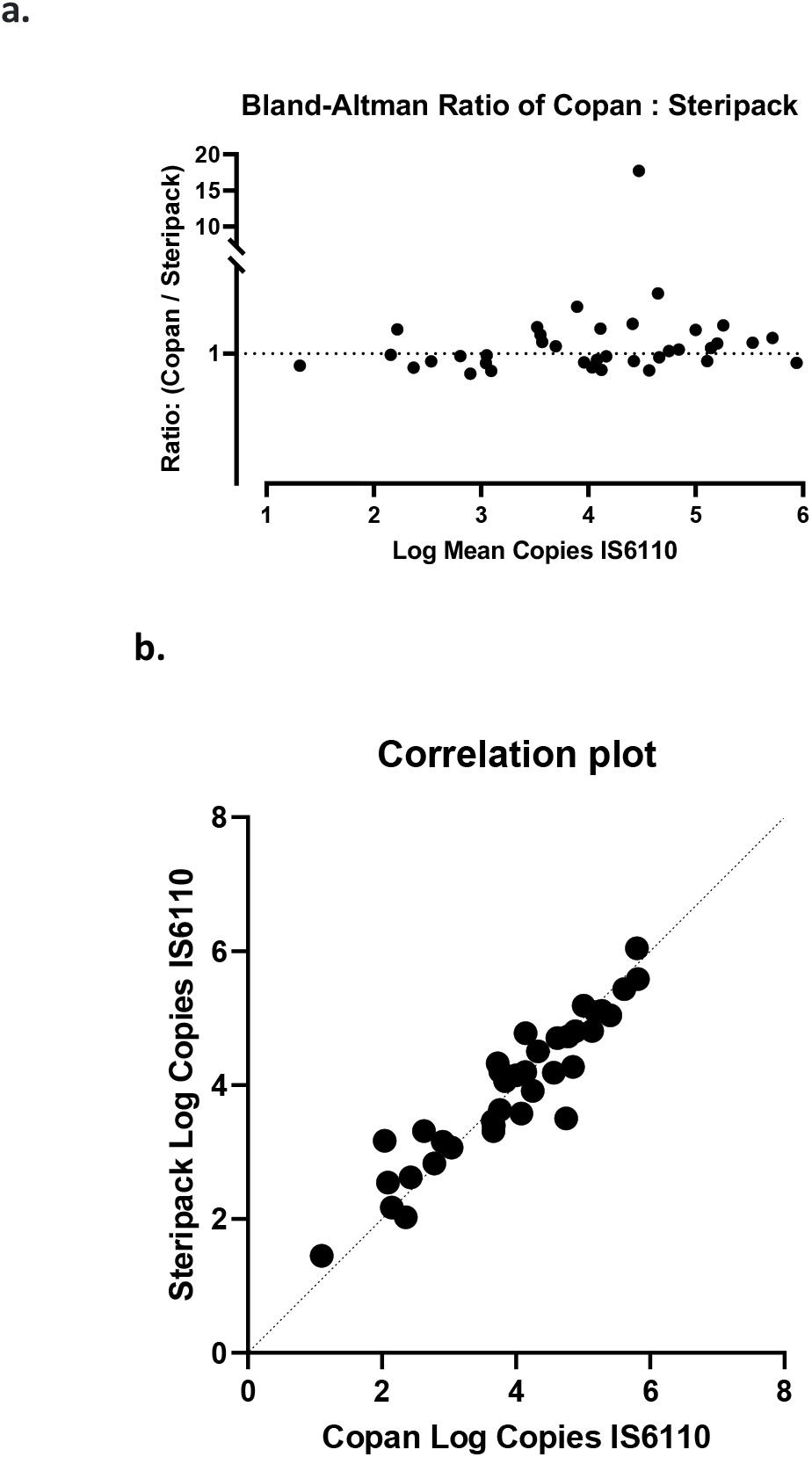
Recovery of IS6110 copies from Copan vs. Steripack swabs. **a**. Bland-Altman ratio of measured IS6110 copies recovered by Copan vs. Steripack divided by log mean measured copies of IS6110. **b**. Measured log copies 1S6110 recovered from Steripack vs. Copan.

## DISCUSSION

We created and evaluated a highly sensitive tongue swab qPCR method to better understand the limits of performance for tongue swab-based molecular assays under “best-case-scenario” circumstances. Overall sensitivity and specificity were nearly equivalent to sputum Xpert Ultra when compared to MRS, suggesting MTB is typically present on the tongues of people with pulmonary TB. However, participants in this study presented with TB symptoms, and of Xpert Ultra-positive participants, 93% had sputum bacillary load categorized as “Low” or higher. Further testing of participants with “Very Low” and “Trace” results must be conducted to understand tongue swab yield for these groups.

For participants with measurable MTB on tongue swabs, qPCR revealed that most had quantities sufficient for detection with less-sensitive methods. Simple math suggests one may be able to decrease test input volume, complexity of the lysis instrument, or use a more common polymerase and still have detectable quantities of MTB, making tongue swabs a reasonable specimen type even for point-of-care platforms.

We also demonstrated that expensive and laborious DNA extraction steps may be removed from swab-based TB diagnostic workflows, decreasing cost, consumables, and waste, reducing user steps and TAT, and minimizing contamination risks associated with these procedures. We determined that heating 10 minutes is sufficient for inactivation of nucleases is mycobactericidal **(Supplement 3)**. Our findings also underscore the importance of efficient TB lysis methods. There are currently few MTB lysis tools amenable to POC settings, and we emphasize the need for low-cost lysis devices to complement molecular assays.

Encouragingly, serial swabbing results demonstrated that additional MTB may be recovered from the tongue with more sensitive sampling tools, since four of four swabs produced positive results for all but one participant. Studying yield from two swab types produced two key conclusions: there is flexibility in the type of swab that may be used for sampling, but we are leaving valuable MTB targets on the tongue. For these reasons, we believe swab design innovations may increase sensitivity. Our early prototyping with 3D-printed “plastic swabs” did not produce better sampling efficiency than Copan (data not shown), but refinements to surface chemistry (e.g., flocking) and form (e.g., scraping stringency or surface area) may increase performance.

Serial swabbing results should also serve as a reminder to take caution when designing multi-swab studies to compare variables, as each swab is likely to yield varying amounts of MTB targets. We suggest randomization of swab order or collecting timed, half-tongue swabs when smaller sample sizes are desired, and we demonstrated the efficacy of this approach with our study design. While we swabbed each participant for 30 seconds to ensure uniformity between swabs during two of three sub-studies, we confirmed that 15 seconds of total swab time is adequate to saturate the swab, based on results from our third sub-study.

The present findings provide important research tools and demonstrate the feasibility of same-day molecular testing of tongue swabs, but findings are limited to two clinics, and expanding to multiple sites and geographies is a top priority. Performance must still be assessed in the groups who may benefit the most from non-sputum sampling options, such as PLHIV, children, and household contacts of index cases. However, our results reinforce the efficacy of tongue swabs for TB diagnosis and unlock keys to developing a class of highly sensitive novel non-sputum tests.

## Supporting information

Supplemental document 1: Oligo sequences

Supplemental document 2: Tongue swab collection, processing, and qPCR SOP

Supplemental document 3: MTB H37Rv culture, contrived sample generation, and lysis techniques

Supplemental document 4: Nuclease inactivation and biosafety experiments

Supplemental document 5: Data from combined studies

## Data Availability

All data produced in the present study are available upon reasonable request to the authors

## ACKNOWLEDGEMENTS

Research reported in this publication was supported by funding from the National Institute of Allergy and Infectious Diseases of the National Institutes of Health under award number U01AI152087, and from Global Health Labs. The authors would like to thank the patients, staff and administration of Mulago National Referral Hospital and Kisenyi Health Centre for their support and participation in the study.

