## Supplemental document 2: Tongue swab collection, processing, and qPCR SOP for "New manual qPCR assay validated on tongue swabs collected and processed in Uganda shows sensitivity that rivals sputum-based molecular TB diagnostics"

### Purpose

This document is intended for use on tongue swab samples collected from people presenting with TB symptoms. These samples will be analyzed for the presence of *Mycobacterium tuberculosis* (MTB) by qPCR. The procedures described are:

1. Lab safety
2. Self-swabbing for standard curves
3. Swab elution
4. Sample heating
5. Sample bead beating lysis
6. GHL MTB qPCR
7. Operation of PCR machine
8. Data analysis

### Scope

This SOP applies specifically to the evaluation of tongue swabs from people >=16 years of age with history of TB symptoms. The procedure as presented is not validated by a regulatory body for clinical diagnostics; as such, qPCR results obtained through its application should not be used to direct decisions about individual participant treatments. Samples will be collected by clinic staff in accordance with the SOP entitled "Tongue Swab Sample Collection and Storage."

#### 1. Biosafety

1.1. Carefully follow all health and safety regulations according to your institution's policies.

#### 2. Tongue swab collection

After participant has been enrolled, select the following materials (described **Table 2** below). Ensure that you have tube racks available for sample storage.

**For each subject sample collection**:

- The subject should not brush their teeth or use mouthwash for at least 30 minutes (minimum) to 1 hour (optimal) before giving the sample.
- Subject should not eat or drink after arriving at the clinic until all swabs have been collected.
- Study staff collecting and processing samples will wear appropriate personal protective equipment (PPE) according to local guidelines for TB and COVID-19. Minimum recommended:
  - Wear recommended duty attire
  - Wear N95 mask during sample collection
  - Wear safety glasses (minimum) or face shield during sample collection
  - Put on fresh pair of gloves while preparing collection materials. Wear gloves during collection. Change gloves between participants.

**NOTE: Collection devices, swab heads, and collection tubes should be considered infectious at all times following sample collection.**

2.1. **Tongue swab collection procedure:**

**Tongue swab sample collection notes:**

- Tongue swab sampling method is derived from https://jcm.asm.org/content/jcm/57/3/e01847-18.full.pdf.
- Subject should close their mouth between sample collections.
- The Copan swabs have a breakpoint 30mm from the swab head. When inserting the swab head into the collection tube, these swab heads will be broken off at the breakpoint.
- Study staff must disinfect hands between patients.

2.1.1. Prefill sterile collection tubes with screw cap and O-ring with sterile, molecular grade 500µl 1X Tris-EDTA pH 8.0 and deliver to clinic where sample collection occurs.
2.1.2. With clean gloves, unscrew cap slightly from collection tube.
2.1.3. Remove swab from packaging. Do not touch collection pad of swab on surfaces other than subject's tongue.
2.1.4. Ask subject to stick out tongue.
2.1.5. Swab the entire length and breadth of the front three quarters of the visible tongue dorsum, focusing of the posterior section of this area and ensuring that firm pressure is applied (enough to see the Copan swab stem flex) and the swab is regularly rolled to saturate the entire swab head. It

should take approximately 15 seconds to thoroughly swab in this manner, and thorough coverage is essential to high quality results.

2.1.6. After swabbing procedure is complete, place head of Copan swab into collection tube, and flex handle to break off swab head into tube.

2.1.7. Securely replace cap on tube containing swab.

#### 3. Air contamination control collection

This air sampling contamination control is to be <u>conducted at each clinic site, at the site where regular participant sample collection normally occurs, one day per week.</u>

3.1. Select one tube labeled “Air Control.”

3.2. Unscrew cap from the tube to the point of being loose, but still attached to tube.

3.3. Open packaging of Copan FLOQswab, and remove swab from packaging, being careful not to touch swab head on surfaces other than as directed in subsequent lines.

3.4. Swirl swab in the air for 15 seconds.

3.5. Place the swab head in the tube with the breakpoint positioned on the tube edge. Flex the swab handle to break the swab head into the tube.

3.6. Secure the cap of the tube.

#### 4. Post-collection sample handling:

4.1. For all samples, including the “air sampling contamination control” samples, immediately following sample collection, place all tubes in lidded box containing cold packs.

4.2. At end of collection day, clean interior of clear plastic lidded box (with wipe of 70% ethanol or isopropanol.)

4.3. Transfer samples to lab for same-day processing or storage at -80°C.

#### 5. Collection of self-tongue-swab for standard curves

5.1. Select five Copan FLOQswabs and five 2.0ml screw cap tubes and place in a tube holder.

5.2. Slightly unscrew cap of one of the tubes.

5.3. Carefully remove Copan swab from packaging (be sure not to touch the tip of the swab to anything).

5.4. Swab your tongue for approximately 15-20 seconds.

5.5. Break off swab head into the 2.0ml screw cap tube and recap the tube.

5.6. Repeat the procedure for a total of five swabs.

5.7. Process tubes containing swab heads alongside the clinical samples.

#### 6. Sample Heating

*The first step in the entire process will be to heat the samples, even if they will be stored for processing at another time.*

6.1. **Adding TE and heating tongue swab samples**

6.1.1. Turn on biosafety cabinet (BSC) and make sure fan is running for at least 15 minutes prior to beginning work.
6.1.2. Preheat "dry bath" heater containing heat blocks to 95°C.
6.1.3. Place sample tubes containing swab head in a rack in the biosafety cabinet.
6.1.4. Place an aliquot of TE buffer pH 8.0 in the biosafety cabinet.
6.1.5. Carefully unscrew the lid of the first sample.
6.1.6. Add 500 µl of TE pH 8.0 to the tube containing swab head and replace cap.
6.1.7. Repeat the procedure in steps 6.1.5-6.1.6 until all tubes containing swabs have TE buffer.
6.1.8. Ensure all tube caps are tight, then vortex all tubes containing buffer on highest setting for 15 seconds.
6.1.9. Place tubes in the dry bath heater and heat tubes containing Copan swab head for 10 minutes at 95°C. Allow samples to passively cool for 10 minutes before further processing.
*NOTE: You may perform section 7.1 to prepare bead tubes during heat step.*
6.1.10. After cooling, again vortex all tubes in buffer on highest setting for 15 seconds.
6.1.11. Place tube in minicentrifuge and briefly spin.

#### 7. Sample Bead Beating Lysis

7.1. **Preparing bead tubes**

7.1.1. Carefully remove the lid of a 2ml screw top tube.
7.1.2. Place 2 ml screw top tube (without lid) on weigh balance and tare. The screen should now read "0."
7.1.3. Add RPI 0.1mm glass beads until weigh balance reads "150 mg" or "0.15 g" (a range of 140 to 200mg is acceptable).
*NOTE: These "bead tubes" can be prepared ahead of time as the process is quite tedious. You must re-tare each tube, as there is a variation in the weight of the tubes.*

7.2. **Traditional bead beating**

7.2.1. Place bead tubes in the biosafety cabinet.
7.2.2. Carefully unscrew the lid of the first tube.
7.2.3. Remove entire volume of eluted sample in TE buffer (~380 µl) into tube containing beads and labeled with patient ID and aliquot number.
7.2.4. Screw cap back on tube containing Copan FLOQswab head (all supernatant has been removed) and dispose in the biohazardous waste in the biosafety cabinet.
7.2.5. Repeat steps 7.2.2-7.2.4 for each sample.
7.2.6. Open lid to the Bead Beater and unscrew metal screw (while pulling out the pin). Remove the safety lid and the tube holder cassette.

- Place bead tubes in the cassette in a balanced manner (similar to balancing centrifuge).
- The minimum total of 4 tubes should be opposite one another in a "cross" pattern. This ensures stability of the cassette and lid during the beating process.

7.2.6.1. Place tube cassette in bead beater with lid cover. Pay special attention to the large pin that aligns these two components.
7.2.6.2. Screw the metal holder over the cassette lid as tight as possible and MAKE SURE THE LOCKING PIN IS FULLY COMPRESSED AND ENGAGED. The locking pin will make a clicking sound as the screw is tightened.

7.2.6.3. Close the apparatus and set it to 1 minute. Set an external timer and do three, one-minute (3 x 1min) beating cycles with 1-minute rests in between.
7.2.6.4. Remove sample tubes when procedure is complete. Take caution when unscrewing the metal screw. This can cause a wrist strain when simultaneously pulling out the locking pin.

##### 7.3. Aliquoting bead beaten samples

7.3.1. After bead beating procedure is complete, briefly centrifuge tubes in tabletop microcentrifuge to collect liquid remaining on sides and in lids of tubes.
7.3.2. Remove the entire liquid volume (about 265 µl) into a fresh, appropriately labeled tube.
7.3.3. Place tube with remainder of sample at 4-8°C until ready for GHL qPCR.

#### 8. GHL MTB qPCR

*This portion must take place in PCR Workstation, which is under positive pressure, to avoid contamination of reagents with template.*

- Put on fresh gloves
- Put on clean room lab coat
- Decontaminate PCR workstation with RNAse Away and wipe surface.
- Follow with 70-75% ethanol or isopropanol to remove salts.

##### 8.1. Making qPCR Mix – FOR ONE PLATE OF SAMPLES

*The volumes in this table make enough PCR mix for one plate, or 15 swabs with 5 replicates PCR wells plus standard curves and controls. There will be a small amount of extra volume.*

8.1.1. Make multiplex oligonucleotide (primers and probes) mix by adding **Table 1** contents to a sterile tube.

*Table 1: Oligo pool concentrations*

| Oligo/Reagent Name | Volume | Unit | Final PCR conc. | Starting conc. |
|---|---|---|---|---|
| 01F | 30 | µl | 0.5 µM | 100 µM |
| 01R | 30 | µl | 0.5 µM | 100 µM |
| 103F | 30 | µl | 0.5 µM | 100 µM |
| 103R | 30 | µl | 0.5 µM | 100 µM |
| 01P – FAM (red) | 15 | µl | 0.25 µM | 100 µM |
| 103P – YY (red) | 15 | µl | 0.25 µM | 100 µM |
| RP-F | 4.8 | µl | 0.08 µM | 100 µM |
| RP-R | 4.8 | µl | 0.08 µM | 100 µM |
| RP-P – Cy5 (blue) | 2.4 | µl | 0.04 µM | 100 µM |
| 10X Kapa3G master mix | 600 | µl | 1X | 10X |
| Molecular grade water | 238 | µl | | |
| **Total Volume** | **1000** | **µl** | | |

8.1.2. In PCR Workstation, add 10 µl master mix from steps 5.1.1. to each well of a PCR plate according to the plate map in ***Figure 1.***

| | 1 | 2 | 3 | 4 | 5 | 6 | 7 | 8 | 9 | 10 | 11 | 12 |
|---|---|---|---|---|---|---|---|---|---|---|---|---|
| A | Standard MTB 100000 | Standard MTB 100000 | Sample 1 | Sample 1 | Sample 1 | Sample 1 | Sample 1 | Sample 9 | Sample 9 | Sample 9 | Sample 9 | Sample 9 |
| B | Standard MTB 10000 | Standard MTB 10000 | Sample 2 | Sample 2 | Sample 2 | Sample 2 | Sample 2 | Sample 10 | Sample 10 | Sample 10 | Sample 10 | Sample 10 |
| C | Standard MTB 1000 | Standard MTB 1000 | Sample 3 | Sample 3 | Sample 3 | Sample 3 | Sample 3 | Sample 11 | Sample 11 | Sample 11 | Sample 11 | Sample 11 |
| D | Standard MTB 100 | Standard MTB 100 | Sample 4 | Sample 4 | Sample 4 | Sample 4 | Sample 4 | Sample 12 | Sample 12 | Sample 12 | Sample 12 | Sample 12 |
| E | Standard MTB 10 | Standard MTB 10 | Sample 5 | Sample 5 | Sample 5 | Sample 5 | Sample 5 | Sample 13 | Sample 13 | Sample 13 | Sample 13 | Sample 13 |
| F | Standard MTB 1 | Standard MTB 1 | Sample 6 | Sample 6 | Sample 6 | Sample 6 | Sample 6 | Sample 14 | Sample 14 | Sample 14 | Sample 14 | Sample 14 |
| G | Standard MTB 0.1 | Standard MTB 0.1 | Sample 7 | Sample 7 | Sample 7 | Sample 7 | Sample 7 | Sample 15 | Sample 15 | Sample 15 | Sample 15 | Sample 15 |
| H | NTC matrix | NTC matrix | Sample 8 | Sample 8 | Sample 8 | Sample 8 | Sample 8 | ACC | ACC | ACC | ACC | ACC |

*Figure 1: PCR plate map*

8.1.3. Seal plate with adhesive plate seal until addition of standard curves and participant specimens.
8.1.4. Briefly centrifuge (about 20 seconds) in plate centrifuge prior to adding samples to plate.

8.2. **Making standard curves for *Mycobacterium tuberculosis H37Rv* quantitative DNA**

8.2.1. Remove tube containing *M. tuberculosis* (MTB) DNA from -80°C and thaw at room temperature.
8.2.2. Ensure stock concentration has been diluted to 2 x $10^5$ genomes/µl
8.2.3. Vortex thoroughly and touch spin.
8.2.4. Perform 10-fold dilution by adding 20 µl of 2 x $10^5$ DNA to 180 µl of molecular biology grade water.
8.2.5. Vortex thoroughly and touch spin.

NOTE: This is now your working tube. Write your initials and the dilution date on the tube. You may save this tube for up to two weeks at 4°C.

8.2.6. Place seven microcentrifuge tubes in a tube rack and label.
8.2.7. Open each tube.
8.2.8. Thoroughly vortex the tongue matrix. Add 180 µl of mixed tongue matrix to all seven tubes, then close all tubes.
8.2.9. Open the tube labeled "1."
8.2.10. Perform 10-fold dilution by adding 20 µl DNA (diluted in step 8.2.4) to 180 µl tongue matrix. Vortex thoroughly and touch spin (very briefly). Concentration is 2 x $10^3$ genomes/µl. **This is the top dilution of the standard curve.**
8.2.11. Perform 10-fold serial dilutions by adding 20 µl to 180 µl matrix, then thoroughly vortex and briefly touch spin. Concentration is 2 x $10^2$ genomes/µl.
8.2.12. Repeat 10-fold dilution process described above five more times, with lowest concentration at 0.002 genomes/µl. This will be seven total standard curve tubes.
8.2.13. Carefully unseal PCR plate in BSC. Vortex each standard curve tube and and add 50 µl of each dilution to appropriate standard curve wells, according to the plate map in **Figure 1**.

8.3. **Adding processed patient samples to the PCR plate**

8.3.1. In biosafety cabinet, thoroughly vortex each sample, then immediately add 50 µl of lysed sample to the appropriate well on PCR plate containing master mix and standard curves according to the plate maps shown in **Figure 1.** Avoid bubbles.

8.3.2. Add 50 µl of molecular biology grade water to NTC wells (H1 and H2).

8.3.3. Seal plate thoroughly with PCR plate seal.

8.3.4. Place plate in plate centrifuge and spin briefly (about 20 seconds). Remove plate and place in PCR machine.

8.3.5. Place a compression pad on top of the PCR plate to help prevent evaporation.

#### 9. Operation of PCR Machine

9.1. **Setting up software**

9.1.1. Ensure QuantStudio 5 machine has been turned on.

9.1.2. On associated computer, open new file in "QuantStudio Design & Analysis Software."

9.2. **Confirming settings in QuantStudio Design & Analysis Software**

9.2.1. On "Properties" screen, enter the date and PCR plate number as the "Name"

9.2.2. Enter your name for the "User Name" ("Barcode" may be left blank)

9.2.3. Go to the File Menu and Select "Save As," and save to folder on Desktop.

9.2.4. Ensure the following settings are selected:

9.2.4.1. Select "QuantStudio 5 System" from "Instrument type" drop down menu

9.2.4.2. Select "96-Well 0.2-mL Block" from "Block type" drop down menu

9.2.4.3. Select "Standard Curve" from "Experiment type" drop down menu

9.2.4.4. Select "TaqMan Reagents" from "Chemistry" drop down menu

9.2.4.5. Ensure all fields are entered as shown in **Figure 2.**

9.2.4.6. Click "Next" to proceed to the "Method" tab

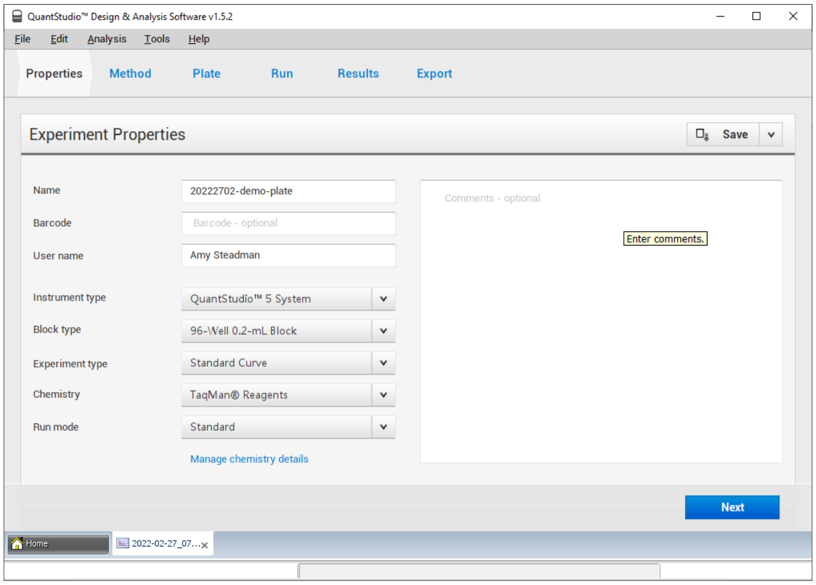

*Figure 2: Properties tab on QuantStudio 5 software*

9.2.5. On the "Method" tab, make sure "Volume" is set to to 60 µl. (Cover at 105°C default temperature)

9.2.6.Program the Experiment Method with the PCR cycling conditions shown in **Table 4.**

9.2.6.1. **Hold Stage** should be set to 98°C for 3:00 minutes (2.5 °C/s ramp rate). This is the initial denaturation step.

9.2.6.2. **PCR Stage Step 1** should be set to 98°C for 5 seconds (2.5 °C/s ramp rate). This is the denaturation cycle.

9.2.6.3. **PCR Stage Step 2** should be set to 64°C for 20 seconds (2.5 °C/s ramp rate). This is the annealing step.

9.2.6.4. **PCR Stage Step 3** should be set to 72°C for 30 seconds (2.5 °C/s ramp rate). This is the extension step. **The camera icon should be clicked on (dark blue) for this step. This enables a fluorescence read after extension.**

9.2.6.5. After PCR Stages have been entered, add a second **Hold Stage** and set to 72°C for 1:00 minute. This is the final elongation step.

9.2.6.6. At the bottom of the **PCR Stage** box, type **45**x into the box for 45 cycles of the PCR stage.

*Table 4: PCR cycling conditions*

| | **Hold Stage Step** | **Temp** | **Time** | **Ramp Rate** | |
|---|---|---|---|---|---|
| Step 1 | Initial denaturation | 98°C | 3:00 | 2.5 °C/s | |
| | **PCR Stage Step (cycling)** | | | | |
| Step 1 | Denaturation | 98°C | 0:05 | 2.5 °C/s | |
| Step 2 | Annealing | 64°C | 0:20 | 2.5 °C/s | |
| Step 3 | Elongation | 72°C | 0:30 | 2.5 °C/s | fluorescence read |
| | **Repeat x 45 cycles** | | | | |
| | **Hold Stage Step** | | | | |
| | Final elongation | 72°C | 1:00 | 2.5 °C/s | |
| | **Repeat x 45 cycles** | | | | |

9.2.6.7. Once programming the Method is complete and matches **Figure 4**, click “Next” to proceed to the “Plate” tab.

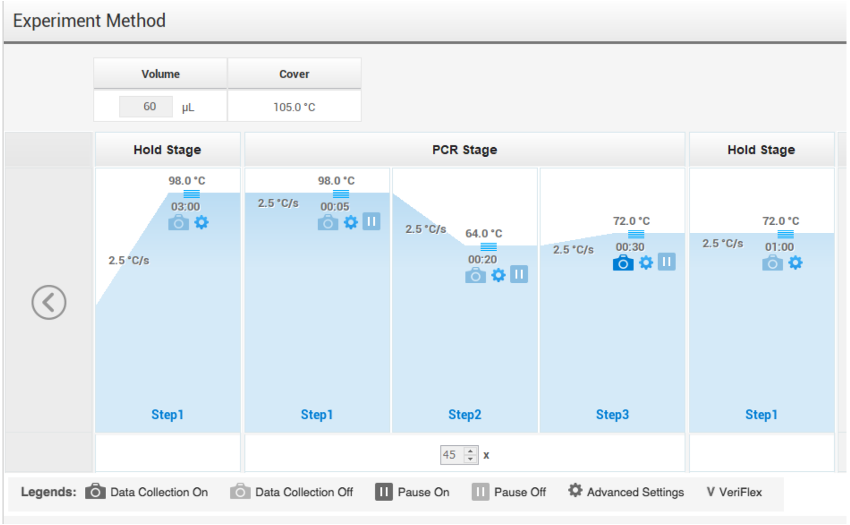

*Figure 3: Thermal cycling conditions in the Experiment Method tab.*

9.2.7. Edit plate set up in the **Plate tab.**

9.2.7.1. In **Advanced Setup** window, add Targets by pressing the **+Add** button.

9.2.7.1.1. Name the first target "MTB IS6110," choose "FAM" for Reporter and "NFQ-MGB" for Quencher.

9.2.7.1.2. Name the second target "MTB IS1081," choose "VIC" for Reporter, and "NFQ-MGB" for Quencher.

9.2.7.1.3. Name the third target "RNaseP," choose "Cy5" for Reporter and "NFQ-MGB" for Quencher.

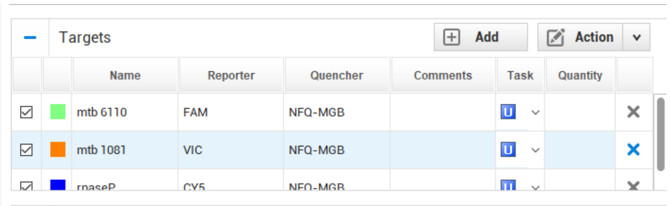

*Figure 4: Assigning Targets in QuantStudio software*

9.2.7.2. Return to **Quick Setup** tab, and add standard curves, by right clicking, then selecting "Define and Set up Standards" from drop down menu.

9.2.7.2.1. Under the **Select a target** section, change "Model" to "Multiplex" and select "MTB" as the target for this standard curve.

9.2.7.2.2. In the **Define the standard curve** section, change "# of Replicates" to "2," change "Starting Quantity" to "100000," and change "Serial Factor" to "1:10."

9.2.7.2.3. In the **Select and arrange wells for the standards** section, choose the radio button to arrange standards in rows, and choose "Let Me Select Wells." Select wells A1-F1 and A2-F2 for the MTB standards and click "Apply."

9.2.7.2.4. To finish, click "Close."

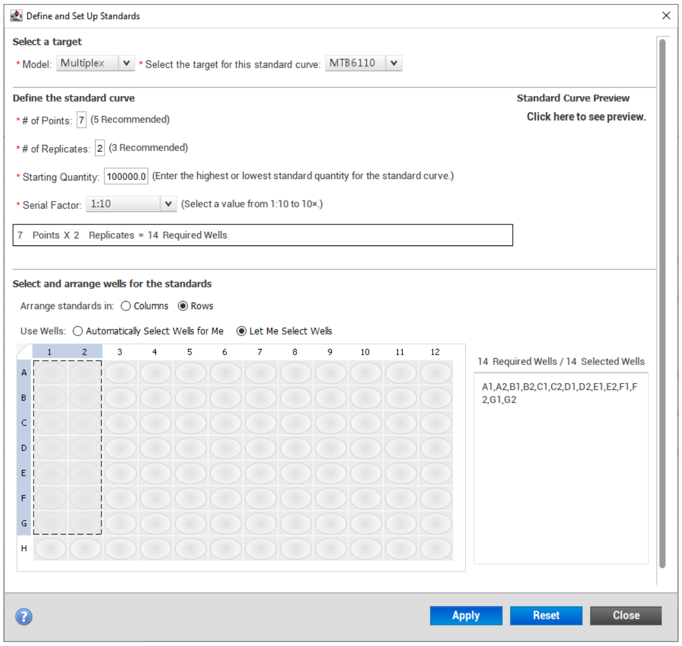

*Figure 5: Defining standard curves*

9.2.7.3. Return to **Quick Setup** tab, and under **Plate Attributes** at bottom of window, change "Passive Reference" from "ROX" to "None" in the drop-down menu.

9.2.7.3.1. Add "MTB IS6110," 'MTB IS1081," and "RNaseP" under **Targets** field.

9.2.7.4. Next, highlight all remaining wells, and assign all targets under the **Target** section on the left. These samples will automatically be categorized as unknowns.

9.2.7.4.1. To assign sample IDs to wells, highlight the wells of interest, then add the ID under "Sample" and hit enter. Multiple wells may be highlighted at once for replicates.

9.2.7.4.2. Continue adding sample IDs to the wells, until all wells have been assigned.

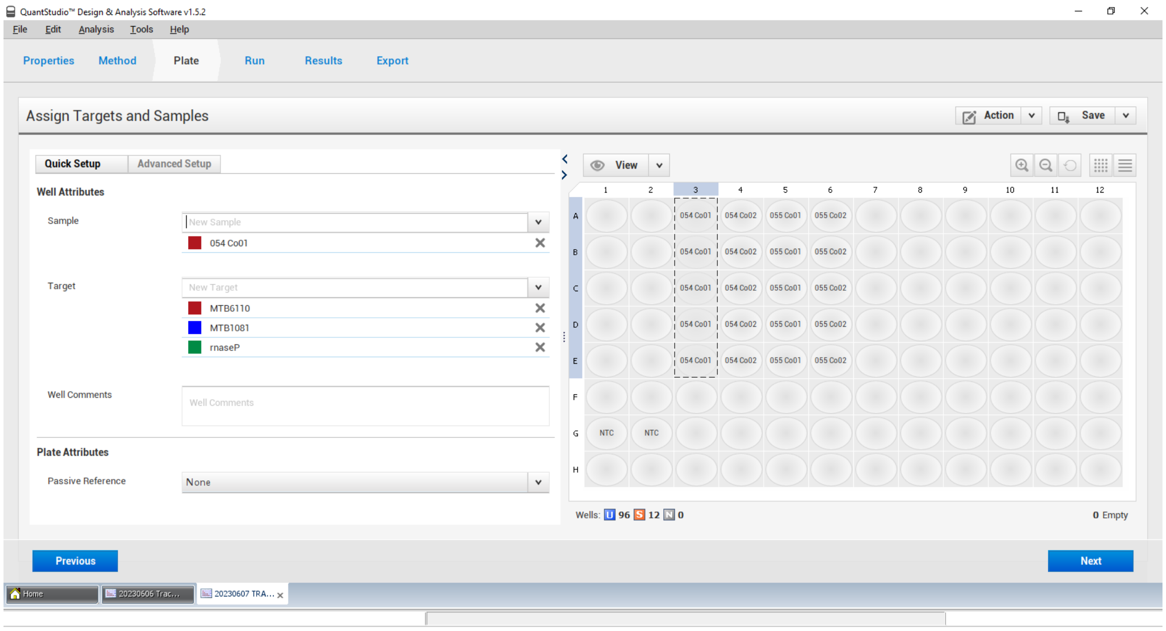

*Figure 6: Assigning sample IDs to wells*

9.2.7.5. Click **Next** at bottom right to proceed to the **Run** tab.

9.2.7.6. Insert plate into PCR machine, by clicking the open drawer icon on the machine.

9.2.7.7. When the drawer opens, place plate in machine and ensure it is evenly seated, then click the drawer icon to close the drawer.

9.2.8. In the **Run** window on the computer, select the blue "Start Run" button, and the PCR machine ID will appear below the button. Click on the machine ID number, and the run will begin. *Note: the run should take about 67 minutes.*

### 10. Data Analysis

10.1. Manually set thresholds to ensure uniformity across PCR plates from different days.

10.1.1. In the "Analysis" menu, choose "Analysis settings."

10.1.2. Click on the first target name, then unclick the "Automatic Threshold" button on the right side of the screen. The box shown in Figure 7 will appear.

10.1.3. In the box entitled "Threshold," change the value to the appropriate threshold setting (note that these values have been empirically determined to work well for this combination of oligos, fluorophore, and the QS5 instruments that have been evaluated:

10.1.3.1. For IS1081, use 15000

10.1.3.2. For IS6110, use 100000

10.1.3.3. For RNaseP, use 10000

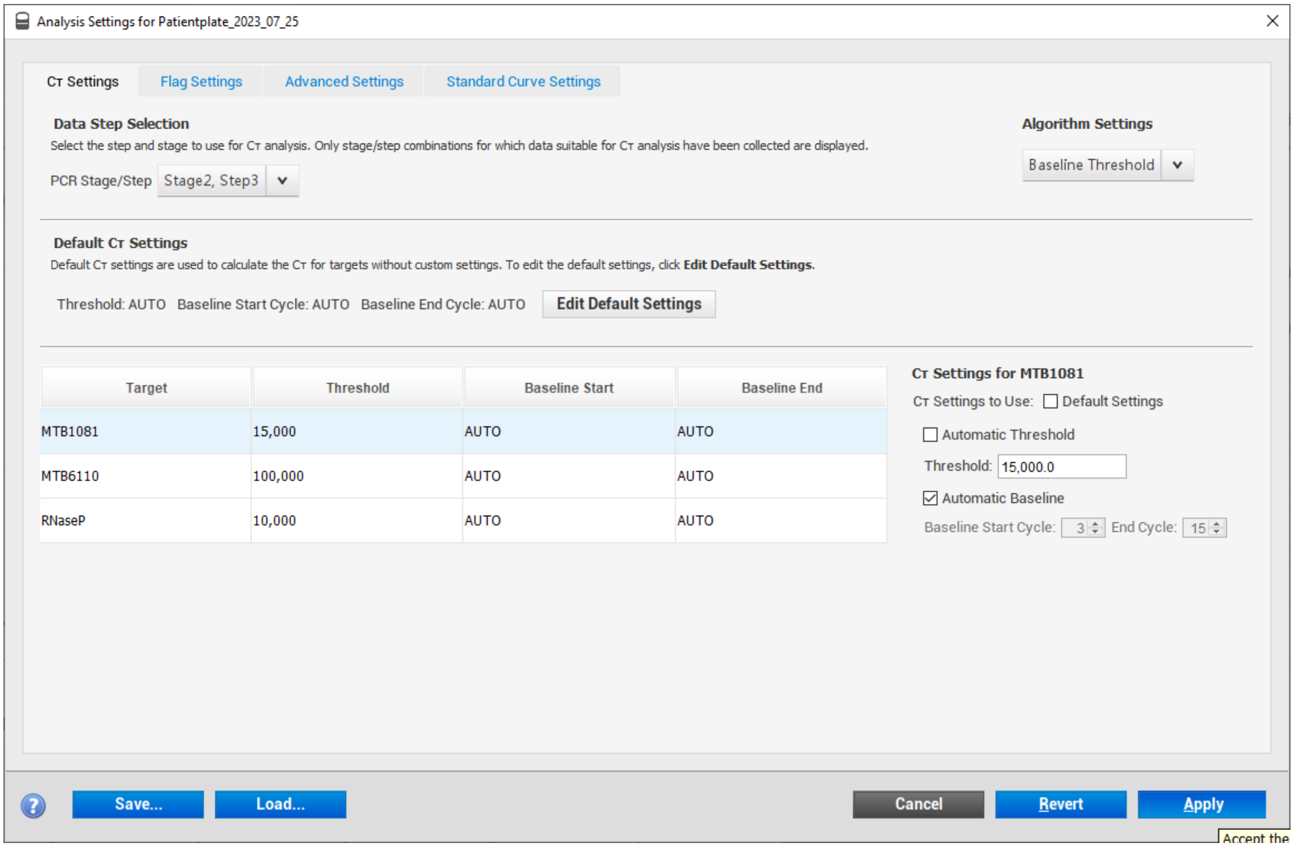

*Figure 7: Changing PCR threshold settings*

10.2. Examine standard curves and ensure that efficiency is between 90-110%.

10.3. Ensure no amplification occurred above the threshold in NTC wells.

10.4. Identify positive patient samples where replicates contained wells with Ct values below 40.

#### 11. Exporting Data

11.1. Upon completion of the run and after changing settings, click **Export** tab at top of window.

11.1.1. Choose **Location** by hitting the "Browse" button and choosing the appropriate folder.

11.1.2. Select **Save** button and choose "Save as" from drop-down menu to save .eds file. Ensure plate date and number are in the file name.

11.1.3. Next choose **Export** button from top right of window to export an Excel .xls file.

### 12. Appendices

**Appendix 1. Abbreviations**

| | |
|---|---|
| BB | Traditional bead beating method |
| BSC | biosafety cabinet |
| GHL | Global Health Labs |
| MTB | *Mycobacterium tuberculosis* |
| mL | milliliter |
| NAAT | nucleic acid amplification testing |
| qPCR | quantitative polymerase chain reaction |
| TB | tuberculosis |
| TE | Tris-EDTA |
| µL | microliter |

**Appendix 2. Equipment and Consumables Required**

| Equipment required | Manufacturer | Catalog number |
|---|---|---|
| Bead beater | BioSpec | |
| Biosafety cabinet class II | Baker SterilGARD | SG404-INT |
| Pipette 100-1000 µl | Eppendorf Research Plus | 3123000063 |
| Pipette 20-200 µl | Eppendorf Research Plus | 3123000055 |
| Pipette 2-20 µl | Eppendorf Research Plus | 3123000918 |
| Pipette 0.2-1.0ul | Eppendorf Research Plus | |
| Multichannel pipette 10-100 µl | Eppendorf Research Plus | |
| Refrigerator (4-8°C) | LG Top Freezer Refrigerator | GN-B202SQBB |
| Freezer (-20°C) | | |
| Deep freezer (-80°C) | | |
| Tube racks | | |
| Weigh balance | VWR | |
| Vortex mixer (2) | | |
| Heat dry bath with blocks | Thermo Scientific | 88870006 |
| Tabletop mini-centrifuge (2) | | |
| PCR Workstation | AirClean Systems | AC648LFUVC |
| Biosafety cabinet class II | Baker SterilGARD | SG404-INT |
| QuantStudio 5 Real-Time PCR system | ThermoFisher Scientific | A28574 |
| **Consumables required** | **Manufacturer** | **Catalog Number** |
| Disposable lab coat | | |
| Safety goggles | | |
| Biohazard bag | | |
| Gloves sterile nitrile | | |
| Copan FLOQSWAB with 30mm breakpoint | Copan | 520CS01 |

| | | |
|---|---|---|
| Sterile storage tubes 2.0 ml screw top | VWR | 76417-214 |
| PREEMPT tuberculocidal cleaning agent | | |
| Sodium hypochlorite [diluted to 0.5-0.6%] | JIK brand [3.5% concentrate] | 330355 |
| Ethanol (neat) | Honeywell [absolute] | 32221 |
| RNase Away cleaning agent | | |
| Microcentrifuge tubes | (Eppendorf or similar) | |
| Sterile filtered pipette tips 1000 long shaft | | |
| Sterile filtered pipette tips 2-200 µl | | |
| Sterile filtered pipette tips 2-20 µl | | |
| 10mM Tris-HCl 1mM EDTA (1X TE) pH 8.0 | Sigma-Aldrich | 93282 |
| 0.1mm glass disruption beads | RPI | 9830 |
| Oligonucleotides (9) | See Supplement 3 for sequences | |
| 01F (IS6110 forward primer) | | |
| 01R (IS6110 reverse primer) | | |
| 103F (IS1081 forward primer) | | |
| 103R (IS1081 reverse primer) | | |
| 01P (IS6110 probe with FAM label) | | |
| 103P (IS1081 probe with Yakima Yellow label) | | |
| RP-F (RNaseP forward primer) | | |
| RP-R (RNaseP reverse primer) | | |
| RP-P (RNaseP probe with Cy5 label) | | |
| Sterile microtubes | | |
| Sterile molecular grade water | VWR | 02-0201-1000 |
| Kapa 3G 10X master mix | Roche | |
| *M. tuberculosis* DNA (standard curve) | custom reagent (contact GHL) | |
| Compression pads for PCR | | |
| MicroAmp EnduraPlate Optical 96-well PCR plate | Applied Biosystems | 4483354 |
| Plate seals | BioRad | |
