## Supplemental document 3: MTB H37Rv culture, contrived sample generation, and lysis techniques for "New manual qPCR assay validated on tongue swabs collected and processed in Uganda shows sensitivity that rivals sputum-based molecular TB diagnostics"

| Version number | Revision(s) & reason for amendment | Date of release |
| --- | --- | --- |
| 1.0 | Initial release | 22 Nov. 2020 |
| 2.0 | Updates to swab collection time, heating time, and generating spiked swabs using tongue matrix, saliva, and MTB culture | 21 June 2023 |
| 3.0 | Changed expected cell density in section 1.4.5 | 22 June 2023 |

### Purpose

This document is intended as a guide for generating MTB cell culture and contrived oral swab samples. Methods outlined in the sections below can be used to simulate clinical TB-positive samples.

1. MTB H37Ra culture methods and considerations
2. Collecting, heating, pooling, and spiking oral matrix with MTB culture
3. Collecting tongue matrix and saliva, heating, pooling, and spiking swabs with MTB culture
4. Bead beating lysis of MTB



*Confidential*

### Abbreviations

| | | | | | |
|---|---|---|---|---|---|
| BSC | Biosafety cabinet | GHL | Global Health Labs | LOD | Limit of detection |
| MTB | *Mycobacterium tuberculosis* | mL | milliliter | NAAT | Nucleic acid amplification test |
| TB | tuberculosis | TE | Tris-EDTA | µL | microliter |

### Materials

| **Equipment** | **Manufacturer** | **Catalog number** |
|---|---|---|
| Tube roller | Waverly | TR6E |
| Biosafety cabinet class II | | |
| Pipettes 100-1000 µl | | |
| 1 liter Pyrex bottle | | |
| Autoclave | | |
| Refrigerator (4-8°C) | | |
| Tube rack | | |
| Vortex mixer | | |
| 37°C incubator capable of housing tube roller | | |
| Magnetic stir bar | | |
| Stirring hot plate | | |
| Dry bath heater | ThermoFisher Scientific | 88870008 |
| Heat block for dry bath heater | ThermoFisher Scientific | 88870112 |
| Biospec Mini Bead Beater 16 | Biospec | 607 or 607EU |

| **Consumables** | **Manufacturer/Vendor** | **Catalog Number** |
|---|---|---|
| Disposable lab coat, cuffed and liquid resistant | | |
| Biohazard bag | | |
| Gloves sterile nitrile | | |
| 50 ml conical tubes | Thermo Scientific or similar | 339653 |
| PREempt Plus tuberculocidal disinfectant | Contec, Inc. | 2B101 |
| Ethanol (neat) | | |
| Sterile storage tubes 2.0ml screw top | VWR | 76417-214 |
| Sterile filtered pipette tips | | |
| MTB H37Ra | ATCC | 25177 |
| 20% Tween 80 | | |
| 50% Glycerol | | |
| Middlebrook 7H9 Broth Base | Sigma-Aldrich | M0178-500G |
| Middlebrook ADC Enrichment, 100mL per bottle | VWR | 212352 |
| Copan FLOQswab | Copan | 520CS01 |
| Nuclease-free Tris-EDTA, pH 8.0 (or other elution buffers) | Millipore-Sigma | 8890-100ML |
| 0.1mm glass disruption beads | RPI Corp | 9830 |



*Confidential*

### Methods

#### 1. *Mycobacterium tuberculosis* H37Ra culture

*The following section describes culturing of attenuated (non-virulent) MTB H37Ra. Observe your institution's health and safety protocols when handling MTB culture and human-derived samples.*

##### 1.1. Making liquid media 7H9 with Middlebrook Enrichment, 0.1% Tween 80 and 0.2% Glycerol

**1.1.1.** To an autoclaved 1-liter bottle, add 4.7g 7H9 Middlebrook broth powder to 900 mL $dH_2O$ and a magnetic stir bar. Place flask on stirring hot plate, with heat and stir settings on medium.

**1.1.2.** Cap loosely and mark with autoclave tape. Autoclave contents of bottle at 121°C for 30 minutes.

**1.1.3.** Let liquid cool to room temperature.
*NOTE: THIS IS AN IMPORTANT STEP, DO NOT PROCEED UNTIL THIS STEP IS COMPLETE. REAGENTS IN SUBSEQUENT STEPS WILL DENATURE AT TEMPERATURES ABOVE 55 °C.*

**1.1.4.** Move bottle into BSC (previously wiped down with 70% ethanol to ensure that the next steps are conducted in a sterile environment). Steps after this should be conducted in the BSC.

**1.1.5.** Add 4mL 50% glycerol solution to flask, allow 2 minutes of stirring to elapse such that all contents of the flask are now homogenous.
*NOTE: pre-filter sterilize 50% glycerol in preparation to add to media.*

**1.1.6.** Add 5 mL 20% stock Tween 80 to 7H9 to media. Slowly stir contents of flask. Again, pre-filter sterilize Tween 80 in preparation to add to media.
*NOTE: Tween 80 detergent helps maintain dispersed culture of mycobacteria as well as acting as a carbon source for MTB metabolism. Alternate detergents such as Tyloxopol can be used, however, this cannot be used as a carbon source and as such results in less florid growth of MTB*

**1.1.7.** Add 100mL of Middlebrook ADC Enrichment to 7H9 media. Slowly stir contents of flask.

**1.1.8.** ADC supplement should be used for liquid culture of MTB. For solid culture/plating use Middlebrook OADC supplement.

**1.1.9.** Cap bottle and store 4 °C fridge. Label as '7H9 + GAT (Glycerol + ADC + Tween 80)
*NOTE: Media can be used for up to 3-4 months*

##### 1.2. Inoculating and expanding primary MTB culture

1.2.1. Frozen glycerol stocks used in 1.2.2 were earlier expanded at GH Labs from an ATCC H37Ra stock. Refer to ATCC recommendations for making glycerol stocks from expanded cultures.

1.2.2. Dispense 0.5 mL thawed glycerol stock in 9.5 mL media pre-warmed to RT

1.2.3. Expand in a 50mL conical at 37 °C with rotation for 5-7 days until an $OD_{600}$ of 1.0 – 1.2

1.2.4. Tube <u>MUST</u> horizontally roll at 20 RPM and rock gently in a tube roller such as a Waverly TR6E. Alternately, a ThermoScientific Cel-Gro Tissue Culture Rotator (set at a 10-degree angle) works equivalently. These are just a few examples, as there are alternate manufacturers that make equivalent devices.
*NOTE: IT IS IMPORTANT TO USE THESE TYPES OF ROLLING DEVICES – expanding MTB cells in a traditional bacterial shaker with tubes held upright causes MTB cells to settle at the bottom and clump, which leads to inadequate exposure to oxygen and growth factors in the media. This matters less with cultures such as E. coli, but it is important for MTB culture.*



*Confidential*

1.2.5. The maximum volume of liquid cannot exceed 20% of the total volume of the tube (e.g., 10 mL in a 50 mL conical tube). This ensures adequate aeration of the mycobacterial culture and reduces risk of liquid escape from the tube.

1.2.6. Place at 4 °C to arrest the cells when density reaches an $OD_{600}$ of ~1.0 – 1.2
*NOTE: Time to reach this density depends on quality and starting density of frozen MTB stock, but most typically takes 4 to 6 days.*
*Primary culture stays viable in fridge for up to 6 weeks for seeding secondary cultures. While arrested from active growth, the cells remain viable and intact.*

1.3. **Secondary culture**

1.3.1. Start weekly or as needed (typically on a Thursday to be ready on Monday or Tuesday)
1.3.2. Vigorously vortex primary culture (previously arrested at 4 °C) and seed 150uL in 10mL fresh media
1.3.3. Use the 50mL conical and incubation conditions as described above for the primary culture
1.3.4. Again, arrest at 4°C when density reaches an $OD_{600}$ of ~1.0 – 1.2 after 4-6 days of expansion

**1.4. Prepping cells for experimentation**

*Perform the following steps to remove secondary culture from 4°C, pellet, and replace spent media with fresh supernatant.*

1.4.1. Vortex vigorously (15 seconds on high) and transfer appropriate volume of culture (typically 1-2 mL) and centrifuge at 3000 *x g* for 10 minutes at ambient temperature.

1.4.2. Remove all of supernatant
*NOTE: Pellet may be loose but it is acceptable to lose some cells in order to get rid of all the spent growth media. This removes the majority (99%) of free DNA that can contribute to high background.*

1.4.3. Resuspend in fresh media (typically a lower amount than the original culture volume removed).
*NOTE: It's important to use media in this step because the Tween-80 helps ensure a continued single cell suspension.*

1.4.4. Vortex vigorously.

1.4.5. Adjust to $OD_{600}$ of ~1.0 by the addition of fresh media.
*NOTE: At this density, cells should be approximately 1E+06 to 2E+06 cells/µL. This is based on using GHL highest yielding lysis method to date (heat + bead beating) and downstream analyses by DD-PCR and qPCR using calibrated standards.*

1.4.6. Return the prepped cells to 4°C. They will settle but remain healthy over a period of up to 7 days.
*NOTE: Due to settling, monodispersity of cells diminishes slightly as early as 2-3 days. This can be problematic for LOD studies but otherwise fine for experimentation involving relative differences at higher levels of input. Vortex the prepped cells vigorously (15 seconds on high) before use.*



*Confidential*

#### 2. Procedure for collecting, heating, pooling, and spiking matrix with MTB culture

*The following steps describe self-collecting, pooling, and directly spiking tongue swab matrix. These procedures should be used when the objective is to compare variables downstream of sample elution from a swab. For instance, one should use these steps to pool matrix to compare different lysis devices or NAAT assays. Alternatively, the method for spiking swabs is described in Section 3.*

##### 2.1. Procedure for self-collecting tongue swabs

2.1.1. Ensure subject has consumed nothing by mouth for at least 30 minutes prior to swabbing.
2.1.2. Prefill 2mL screw cap tubes with 500 µl TE buffer.
2.1.3. Set timer for 15 seconds.
2.1.4. Remove swab from packaging. Do not touch collection pad of swab to surfaces other than tongue.
2.1.5. Self-swab the length and breadth of the visible front 3/4 of the tongue, focusing on the posterior dorsum but avoiding gag reflex at rear of the tongue and throat.
2.1.6. Apply firm pressure to swab head during swabbing. Swab shaft will flex slightly, but not bend.
2.1.7. Brush left-right and back-forth, rotating the swab to cover all surfaces during sample collection.
2.1.8. Sample collection should cover length/breadth of area of interest for about 15 seconds.
2.1.9. After collection, insert swab head in to the 2 mL screw cap tube containing 500 µL of TE buffer, and break swab head into tube at 30mm breakpoint.
2.1.10. Secure cap on tube.

##### 2.2. Heating oral swab samples in TE buffer

*Heating serves to inactivate nucleases which may be released during cell lysis. It renders the samples biosafe and will kill (but not lyse) MTB cells.*

2.2.1. Ensure all tube caps are tight, then vortex tube on highest setting for 15 seconds.
2.2.2. Place tubes containing swab heads in standard heat block (Thermo Scientific Cat. # 88870006) and heat at 95°C for 10 minutes to inactivate samples. Allow samples to passively cool for 10 minutes before further processing.
2.2.3. After cooling, vortex tube on high for 15 seconds, flick tube (or briefly spin down in a mini centrifuge) to drive residual liquid down from walls of tube and remove lid.
2.2.4. Transfer all of sample into clean appropriately labeled screw cap tube.
*NOTE: It is usually possible to recover about 380µl from a swab collected into 500µl buffer.*
2.2.5. If pooling, transfer eluent from all tubes into one tube of "pooled tongue matrix" and vortex.

##### 2.3. Spiking contrived oral matrix with MTB

2.3.1. Remove prepared cells from 4 °C (section 1.4.6.).
2.3.2. Vortex vigorously and dilute 1:10 or 1:100 in fresh media.
2.3.2.1. Initial dilution step must be performed in media with Tween 80 to maintain monodispersity and avoid downstream clumping.
2.3.3. Cell suspension can now be further diluted in tongue matrix (generated in section 2.2.5.)


*Confidential*

#### 3. Procedure for collecting, heating, pooling, and spiking swabs with MTB culture

*The following steps describe self-collecting swabs and saliva, pooling, spiking matrix with cells, then dipping tongue swab swabs into resulting mixture for spiked swabs (vs. spiked free matrix). These procedures should be used when objective is to compare swab elution or the entire pathway from swab elution through amplification. For instance, one should use these steps to compare elution efficiency with different buffers.*

##### 3.1. Self-collection of swabs and saliva

3.1.1. Ensure subject has consumed nothing by mouth for at least 30 minutes prior to swabbing.
3.1.2. Set timer for 15 seconds.
3.1.3. Remove swab from packaging. Do not touch collection pad of swab to surfaces other than tongue.
3.1.4. Self-swab the length and breadth of the visible front 3/4 of the tongue, focusing on the posterior dorsum but avoiding gag reflex at rear of the tongue and throat.
3.1.5. Apply firm pressure to swab head during swabbing. Swab shaft will flex slightly, but not bend.
3.1.6. Brush left-right and back-forth, rotating the swab to cover all surfaces during sample collection.
3.1.7. Sample collection should cover length/breadth of area of interest for about 15 seconds.
3.1.8. After collection, insert swab head in to the dry 2mL screw cap tube (containing no buffer), and break swab head into tube at 30mm breakpoint.
3.1.9. Secure cap on tube.
3.1.10. Collect 600µl of saliva from the same donor in a wide 5 ml tube.
*NOTE: You must collect 600µl saliva for each swab collected.*
3.1.11. Pipette 500µl of saliva into the tube containing the tongue swab head (from step 3.1.7.)
3.1.12. Vortex the tube on highest setting for 15 seconds.
3.1.13. Collect the tongue matrix/eluate from the tube and pool if multiple swabs and saliva samples were collected from the same donor to obtain increased amounts of tongue matrix/saliva.
3.1.14. Spike the tongue matrix/saliva with mTB H37Ra such that the final concentration contains the total desired number of cells per reaction in 50µl (see next step).
3.1.15. Add 50µl of the spiked matrix to a 2.0 ml screw cap tube and dip a fresh Copan FLOQswab in the tube. Swirl the swab head well to ensure the matrix gets absorbed on the swab. Once the swab has absorbed the matrix, snap the swab at 30 mm break point. Secure the cap on the lid and label the tube appropriately.
3.1.16. When ready to begin heating and elution process, add 500µl of Tris-EDTA or desired buffer to tube.

##### 3.2. Heating oral swab samples in TE buffer

*Heating serves to inactivate nucleases which may be released during cell lysis. It renders the samples biosafe and will kill (but not lyse) MTB cells.*

3.2.1. Ensure all tube caps are tight, then vortex tube on highest setting for 15 seconds.
3.2.2. Place tubes containing swab heads in standard heat block and heat at 95°C for 10 minutes to inactivate samples. Allow samples to passively cool for 10 minutes before further processing.
3.2.3. After cooling, vortex tube on high for 15 seconds, flick tube (or briefly spin down in a mini centrifuge) to drive residual liquid down from walls of tube and remove lid.
3.2.4. Transfer all of sample into clean appropriately labeled screw cap tube.
*NOTE: It is usually possible to recover about 380µl from a swab collected into 500µl buffer.*


*Confidential*

#### 4. Bead beating lysis of MTB

*Mechanical lysis via bead beating typically generates the highest yield of MTB DNA and serves as our gold standard for comparing other lysis and sample prep methods. We highly recommend the Biospec Mini Beadbeater-16 as a gold standard lysis condition when designing experiments, because it generates the harshest lysis conditions enabling consistency between replicates, regardless of donor-to-donor variation.*

##### 4.1. **Traditional bead beating**

4.1.1. Place 2 ml screw top tube on balance without the screw top cap and tare.
*NOTE: It is imperative to re-tare each tube before adding beads as there is a considerable amount of variation in the weight of the tubes.*

4.1.2. Add RPI 0.1mm glass beads (Cat # 9830) to 150 mg (+/- 10mg).
*NOTE: These "bead tubes" can be prepared well ahead of time as the process is quite tedious.*

4.1.3. Transfer all of the pre-heated sample (~380 µl) to the tube containing 150mg of beads.

4.1.4. Open bead beater lid and unscrew metal screw while pulling out the pin. Remove the safety lid and the tube holder cassette.
- Place bead tubes in the cassette in a balanced manner (similar to balancing centrifuge).
- The minimum total of four tubes should be opposite one another in a "cross" pattern. This ensures stability of the cassette and lid during the beating process.

4.1.5. Place tube cassette in bead beater with lid cover. Pay special attention to the large pin that aligns these two components.

4.1.6. Screw the metal holder over the cassette lid as tight as possible and MAKE SURE THE LOCKING PIN IS FULLY COMPRESSED AND ENGAGED. The locking pin will make a clicking sound as the screw is tightened.

4.1.7. Close the apparatus and set it to one minute. Set an external timer and perform three, one-minute (3 x 1 min) beating cycles with one-minute rests in between.
*NOTE: The one-minute rests are designed to prevent the bead beater from overheating.*

4.1.8. Remove sample tubes when the procedure is complete. Take caution when unscrewing the metal screw. This can cause a wrist strain when simultaneously pulling out the locking pin.

4.1.9. Bead beating will elevate temperature in tubes so let passively cool for five minutes and spin down contents before opening tube for downstream processing.

4.1.10. Samples can proceed directly into NAAT assay.



*Confidential*
