## Supplemental document 4: Nuclease inactivation and biosafety experiments for "New manual qPCR assay validated on tongue swabs collected and processed in Uganda shows sensitivity that rivals sputum-based molecular TB diagnostics"

**Heating for nuclease inactivation**

It is critical to perform a 95°C heat step prior to bead beating (resulting in cell disruption) due to the simultaneous release of nucleases from human cells present in tongue matrix and target DNA from *Mycobacterium tuberculosis* (MTB) cells. In the absence of pre-heating, liberated nucleases will rapidly degrade MTB DNA, reducing the yield by as much as 40-55% within 30 minutes.

Two independent experiments were conducted to evaluate the duration of heating required to inactivate nucleases present in tongue matrix. Briefly, tongue matrix was collected from healthy volunteers as described in the Methods, then eluted matrix was pooled. Cultured MTB H37Ra cells were washed, diluted, and spiked in pooled matrix (5 replicates per condition). The spiked tongue matrix was aliquoted (400 μL) into 2 mL gasketed screw cap tubes containing 150 mg of 0.1mm glass beads. The samples were subjected to heat for 0-, 1-, 2-, 3-, 5-, 10-, 15-, 20-, 25-, or 30-minute duration, then all samples were bead beaten and allowed to remain at ambient temperature for 30 minutes. To inactivate nucleases prior to PCR, all samples were subjected to an additional 10-minute heat step at 95°C, allowed to cool to ambient temperature, briefly centrifuged and added directly to qPCR.

Experiments were performed two separate days to evaluate heat duration from 10 minutes to 30 minutes **(Supplemental Figure 1)**. A second set of experiments was performed on two separate days to evaluate shorter durations from 30 seconds to 5 minutes to establish the minimal heat duration required for heat inactivation **(Supplemental Figure 2)**. Notably the nuclease activity varied among donor pools, so data from the two experiments are shown independently. The experiments confirmed that heating 10 minutes at 95°C is sufficient for nuclease inactivation. Longer time points are unnecessary and heating for less than five minutes results in degradation of target MTB DNA.

Percent Recovery of MTB with Varying Duration of Heat Step

|  | Percent recovery |
| --- | --- |
| Heat Duration (Minutes) 30 | 100 |
| Heat Duration (Minutes) 25 | 95.3546 |
| Heat Duration (Minutes) 20 | 93.3604 |
| Heat Duration (Minutes) 15 | 98.1558 |
| Heat Duration (Minutes) 10 | 99.7568 |
| Controls No pre-heat | 44.5746 |
| Controls Heat alone (no bead beating) | 1.63785 |
| Controls No treatment | 0.00717116 |

**Figure 1.** Mean percent recovery of MTB targets from contrived tongue swab samples subjected to varying duration of heat then bead beaten compared to the gold standard condition of 10 minutes at 95°C plus bead beating.

Percent Recovery of MTB with Varying Duration of Heat Step

|  | % control (10m heat + BB) |
| --- | --- |
| Heat Duration (Minutes) 10 | 100 |
| Heat Duration (Minutes) 5 | 106.141 |
| Heat Duration (Minutes) 3 | 93.4062 |
| Heat Duration (Minutes) 2 | 81.0011 |
| Heat Duration (Minutes) 1 | 69.6035 |
| Heat Duration (Minutes) 0.5 | 64.0634 |
| Controls No pre-heat | 58.9291 |
| Controls Heat alone (no bead beating) | 0.96099 |
| Controls No treatment | 0 |

**Figure 2.** Mean percent recovery of MTB targets from contrived tongue swab samples subjected to varying duration of heat then bead beaten compared to the gold standard condition of 10 minutes at 95°C plus bead beating.

**Heating for biosafety**

MTB H37Ra cells (ATCC 25177) were harvested in exponential phase of growth (OD600 of 1.0), washed, diluted, and subjected to heating for 0, 5, 10, or 15 minutes. After heating, cells were plated at 3.0x10^4^ cells per 10 cm deep fill Middlebrook 7H11 agar plates (N=6 per condition), and counts were confirmed by qPCR. Plates were then incubated at 37°C in sealed, sterile Ziplock bags to prevent desiccation. Colonies for all six plates were per conditions were counted every two weeks. The percentage of survival was determined from the total number of colonies in each heat duration condition relative to the “no heat” condition of approximately 30,000 colonies per plate (1.8x10^5^ per six plates).

| % Survival | No Heat | Heat time course | | |
| --- | --- | --- | --- | --- |
|  |  | 5 min | 10 min | 15 min |
| Week 2 | 100 | 0.035 | 0 | 0 |
| Week 8 | 100 | 0.0472 | 0.0006 | 0 |
